# Epigenetic and transcriptional regulation of osteoclastogenesis in the pathogenesis of skeletal diseases: a protocol for a systematic review

**DOI:** 10.1101/2020.06.05.20123083

**Authors:** Siddhartha Sharma, Anupam Mittal, Aditi Mahajan, Riddhi Gohil

## Abstract

Osteoclastogenesis (OCG) is a multi-stage process that involves formation of activated osteoclasts from bone marrow macrophages. The progression of each stage of OCG is governed by a set of transcription factors and gene regulators, which are genetically and epigenetically regulated at both transcription and post-transcription levels. Epigenetic changes are used to denote interactions between genetic material and environment leading to phenotypic alterations that can be inherited, without any variations in DNA sequence. Epigenetic and transcription regulatory events have profound effects on osteoclast formation and activation; these have been implicated in numerous disorders including osteoporosis, osteopetrosis, rheumatoid arthritis, spondyloarthritis, gout and bone metastasis. We aim to conduct a systematic review to assess possible relationship between key epigenetic and transcriptional regulators of osteoclastogenesis, and their role in specific bone disorders. This is a protocol for the proposed review.

## Background

The dynamic process of bone remodeling is involved in preserving its integrity, and requires active participation of two types of cells – osteoblasts which are involved in bone formation, and osteoclasts which mediate bone resorption. Osteoclasts are myeloid lineage multinucleated giant bone resorbing cells, that are derived from bone marrow macrophages in a process requiring Macrophage Colony Stimulating Factor (M-CSF) and Receptor Activator of Nuclear Factor κB ligand (RANKL) (1). Not only these are required for maintaining bone homeostasis, they also play a pivotal role in skeletal development as well as in the pathogenesis of many bone disorders. One of the key contributing factors in the development of these pathological conditions is the dysregulated activation and maturation of osteoclasts, which results in either excessive or suboptimal bone resorption (2).

Osteoclastogenesis (OCG) is a multi-stage process involving lineage commitment of monocytes to preosteoclasts, followed by their differentiation into committed osteoclasts and finally, their polarization and activation into resorbing osteoclasts (1). The progression of each stage of OCG to the next stage is governed by different transcription factors and gene regulators which are genetically and epigenetically regulated, at both transcription and post-transcription levels (3). Whereas aberrant genetic modifications resulting in gain or loss of functionality of these regulators chiefly account for the pathophysiology of bone disorders, there also occur other forms of regulations which contribute to the disease development, but cannot be explained by genetic inheritance (4). These changes, termed as *epigenetic*, are used to denote interactions between genetic material and environment leading to phenotypic alterations that can be inherited, without any variations in DNA sequence. Epigenetic mechanisms of gene regulation involve chromatin and nucleosome remodeling by means of DNA methylation of cytosine and adenine residues, modifications like acetylation, deacetylation, methylation and phosphorylation of H2/H3/H4 histone proteins and transcriptional or post transcriptional changes by non-coding RNAs such as micro-RNAs (miRNAs), circular RNAs, enhancer RNAs (eRNAs) and long non coding RNAs (lncRNAs) (5). Such mechanisms of gene activation or repression are usually influenced by exogenous factors like age, gender, environment and diet, and are critical not only during developmental processes, but also in the development and progression of various diseased conditions.

## Need for the review

Since epigenetic and transcription regulatory events have profound effects on osteoclast formation and activation, these have been implicated in numerous disorders including osteoporosis, osteopetrosis, rheumatoid arthritis, spondyloarthritis, gout and bone metastasis (6,7). Although there are a number of research publications and a few narrative reviews on this subject, to the best of our knowledge, there is no systematic review on this subject so far.

Hence, we aim to conduct a systematic review to assess possible relationship between key epigenetic and transcriptional regulators of osteoclastogenesis, and their role in specific bone disorders. We believe that this systematic review will help not only to assimilate the emerging research on this fascinating subject, but will also allow us to critically evaluate the available evidence.

## Objectives

The objective of this study is to identify epigenetic and transcriptional factors controlling OCG, that have been shown to play a role in the pathogenesis of skeletal diseases.

## Methods

This systematic review will be conducted in the accordance with the PRISMA guidelines (8).

### Review Protocol

A protocol for the review will be formulated *a priori*, in accordance with the PRISMA-P guidelines (Appendix 1).

### Eligibility Criteria

Studies will be included if they met ***all*** of the following prespecified inclusion criteria:

a. describe epigenetic and/or transcriptional regulation of OCG
b. investigate a specific skeletal disease (or a well-described skeletal disease model)
c. quantify alterations in OCG by any well-described experimental method.

In-vitro, in-vivo and ex-vivo will be considered eligible. There will be no restrictions on the year or language of publication. Since epigenetics is an fast-advancing field, we will accept authors’ definition of epigenetics when considering studies for inclusion or exclusion studies in this review.

The following studies will be excluded:

a. *in-silico* studies
b. studies on cells other than osteoclasts or disorders other than skeletal disorders
c. studies evaluating general mechanisms of osteolysis (rather than specific skeletal disorders)
d. studies on dysfunction in mature osteoclasts rather than OCG
e. studies looking at genetic mechanisms of OCG, rather than epigenetic and/or transcriptional regulation
f. studies looking at post-translational modifications
g. studies that do not quantify OCG by experimental methods
h. review articles, case reports, expert opinions and conference abstracts *Information Sources & Literature Search*

The primary search will conducted by two independent authors on the PubMed and the EMBASE databases using a well-defined search strategy (Table 1). A secondary search will be conducted by screening the references of included articles and as relevant review articles on the subject. Search results will be imported into a reference management software and deduplicated.

**Table 1:**
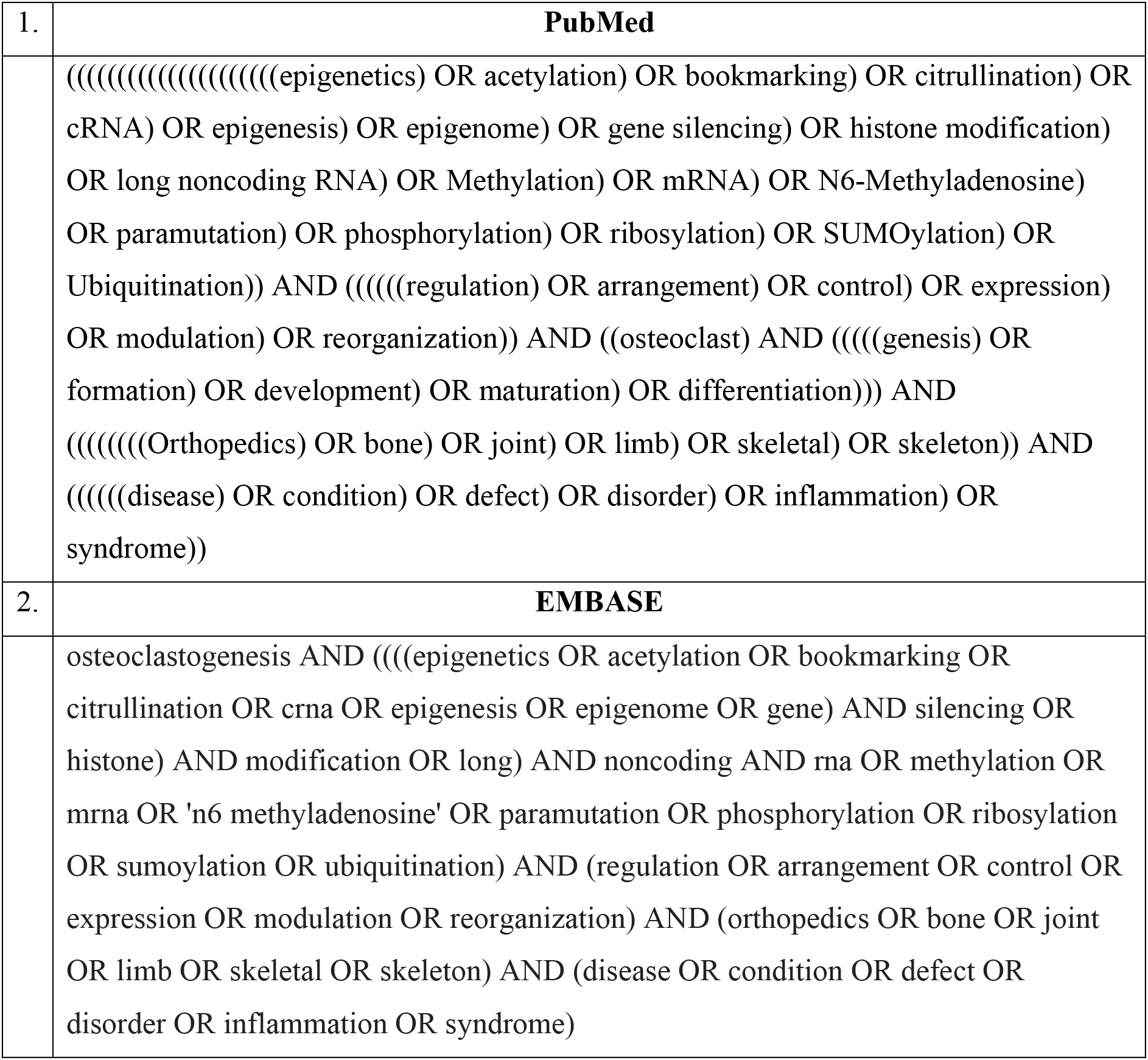
Search Strategy.

### Study Selection

Deduplicated records from the primary search strategy will be screened searched independently by a minimum of two authors. Studies will be included if they meet the inclusion criteria outlined above. Reasons for exclusion of studies for which full-text is obtained will be documented. Conflicts on whether or not to include a particular study in the review will be resolved by mutual consensus.

### Data Collection & Data Items

Data will collected independently on pre-piloted data collection forms by a minimum of two authors. Baseline data items that will be extracted for each study included

a. authors name and year of publication
b. journal
c. study design
d. disease or (disease model) evaluated
e. key experiments performed
f. method(s) of evaluation of OCG.

### Outcome measures

The following outcome measures will be documented:

a. epigenetic or transcriptional regulation factor evaluated
b. the stage of OCG at which the epigenetic/transcription factor acts
c. the mechanism by which the epigenetic/transcription factor acts
d. level of epigenetic/transcription factor in diseased state
e. net effect of the epigenetic/transcription factor on OCG

### Data Analysis & Synthesis of Results

Since we expect to find a diverse range of experimental studies pertaining to our study objectives, only qualitative analysis by narrative synthesis has been planned at the protocol stage. All the prespecified baseline data items and outcome measures will be presented by appropriate tables, figures and diagrams etc.

### Risk of Bias

Risk of bias for in-vivo studies will be determined by a previously described modification (9) of the CAMARADES tool (10). Assessment will be done by two researchers independently and discrepancies will be resolved by mutual consensus. The modified CAMARADES tool (9) consists of the following seven items:

a. publication in a peer-reviewed journal
b. random allocation
c. allocation concealment
d. blinding of personnel assessing the outcomes
e. reporting of sample size calculation
f. compliance with animal welfare organization
g. statement on potential conflicts of interest

Each item of the tool will be graded as ‘*high-risk*’, ‘*low-risk*’ or ‘*unclear risk*’ based on the authors’ judgment.

The assessment of overall strength of evidence will be based on the quality grading of the individual studies in the review.

## Data Availability

Since this is a protocol for systematic review, it does not contain data. However, the search strategy has been presented in Table 1.

**APPENDIX 1:** PRISMA-P (Preferred Reporting Items for Systematic review and Meta-Analysis Protocols) 2015 checklist: recommended items to address in a systematic review protocol*.

| Section and topic | Item No | Checklist item | Page Number |
| --- | --- | --- | --- |
| **ADMINISTRATIVE INFORMATION** |  |  |  |
| Title: |  |  |  |
| Identification | 1a | Identify the report as a protocol of a systematic review | 1,2 |
| Update | 1b | If the protocol is for an update of a previous systematic review, identify as such | NA |
| Registration | 2 | If registered, provide the name of the registry (such as PROSPERO) and registration number | NA |
| Authors: |  |  |  |
| Contact | 3a | Provide name, institutional affiliation, e-mail address of all protocol authors; provide physical mailing address of corresponding author | 1 |
| Contributions | 3b | Describe contributions of protocol authors and identify the guarantor of the review | 1 |
| Amendments | 4 | If the protocol represents an amendment of a previously completed or published protocol, identify as such and list changes; otherwise, state plan for documenting important protocol amendments | NA |
| Support: |  |  |  |
| Sources | 5a | Indicate sources of financial or other support for the review | 1 |
| Sponsor | 5b | Provide name for the review funder and/or sponsor | 1 |
| Role of sponsor or funder | 5c | Describe roles of funder(s), sponsor(s), and/or institution(s), if any, in developing the protocol | NA |
| **INTRODUCTION** |  |  |  |
| Rationale | 6 | Describe the rationale for the review in the context of what is already known | 3,4 |
| Objectives | 7 | Provide an explicit statement of the question(s) the review will address with reference to participants, interventions, comparators, and outcomes (PICO) | 4 |
| **METHODS** |  |  |  |
| Eligibility criteria | 8 | Specify the study characteristics (such as PICO, study design, setting, time frame) and report characteristics (such as years considered, language, publication status) to be used as criteria for eligibility for the review | 5 |
| Information sources | 9 | Describe all intended information sources (such as electronic databases, contact with study authors, trial registers or other grey literature sources) with planned dates of coverage | 5, 6 |
| Search strategy | 10 | Present draft of search strategy to be used for at least one electronic database, including planned limits, such that it could be repeated | Table 1 |
| Study records: |  |  |  |
| Data management | 11a | Describe the mechanism(s) that will be used to manage records and data throughout the review | 6 |
| Selection process | 11b | State the process that will be used for selecting studies (such as two independent reviewers) through each phase of the review (that is, screening, eligibility and inclusion in meta-analysis) | 6 |
| Data collection process | 11c | Describe planned method of extracting data from reports (such as piloting forms, done independently, in duplicate), any processes for obtaining and confirming data from investigators | 6 |
| Data items | 12 | List and define all variables for which data will be sought (such as PICO items, funding sources), any pre-planned data assumptions and simplifications | 6 |
| Outcomes and prioritization | 13 | List and define all outcomes for which data will be sought, including prioritization of main and additional outcomes, with rationale | 6 |
| Risk of bias in individual studies | 14 | Describe anticipated methods for assessing risk of bias of individual studies, including whether this will be done at the outcome or study level, or both; state how this information will be used in data synthesis | 7 |
| Data synthesis | 15a | Describe criteria under which study data will be quantitatively synthesised | Not planned. Quantitative synthesis only. Page 7 |
| | 15b | If data are appropriate for quantitative synthesis, describe planned summary measures, methods of handling data and methods of combining data from studies, including any planned exploration of consistency (such as $I^2$, Kendall's $\tau$) | NA |
|  | 15c | Describe any proposed additional analyses (such as sensitivity or subgroup analyses, meta-regression) | NA |
|  | 15d | If quantitative synthesis is not appropriate, describe the type of summary planned | NA |
| Meta-bias(es) | 16 | Specify any planned assessment of meta-bias(es) (such as publication bias across studies, selective reporting within studies) | NA |
| Confidence in cumulative evidence | 17 | Describe how the strength of the body of evidence will be assessed (such as GRADE) | 7 |

**\* It is strongly recommended that this checklist be read in conjunction with the PRISMA-P Explanation and Elaboration (cite when available) for important clarification on the items. Amendments to a review protocol should be tracked and dated. The copyright for PRISMA-P (including checklist) is held by the PRISMA-P Group and is distributed under a Creative Commons Attribution Licence 4.0.**
*From: Shamseer L, Moher D, Clarke M, Ghersi D, Liberati A, Petticrew M, Shekelle P, Stewart L, PRISMA-P Group. Preferred reporting items for systematic review and meta-analysis protocols (PRISMA-P) 2015: elaboration and explanation. BMJ. 2015 Jan 2;349(jan02 1):g7647.*

## References

1. Boyle WJ, Simonet WS, Lacey DL. Osteoclast differentiation and activation. Vol. 423, Nature. Nature Publishing Group; 2003. p. 337–42.

2. Bi H, Chen X, Gao S, Yu X, Xiao J, Zhang B, et al. Key triggers of osteoclast-related diseases and available strategies for targeted therapies: A review. Vol. 4, Frontiers in Medicine. Frontiers Media S.A.; 2017.

3. Choi Y, Faccio R, Teitelbaum SL, Takayanagi H. Osteoclast Biology: Regulation of Formation and Function. In: Osteoimmunology: Interactions of the Immune and Skeletal Systems: Second Edition. Elsevier Inc.; 2016. p. 41–70.

4. Marini F, Cianferotti L, Brandi ML. Epigenetic mechanisms in bone biology and osteoporosis: Can they drive therapeutic choices? Vol. 17, International Journal of Molecular Sciences. MDPI AG; 2016.

5. Park-Min KH. Epigenetic regulation of bone cells. Connect Tissue Res. 2017 Jan 2;58(1):76–89.

6. Ghayor C, Weber FE. Epigenetic regulation of bone remodeling and its impacts in osteoporosis. Vol. 17, International Journal of Molecular Sciences. MDPI AG; 2016.

7. del Real A, Riancho-Zarrabeitia L, López-Delgado L, Riancho JA. Epigenetics of Skeletal Diseases. Vol. 16, Current Osteoporosis Reports. Current Medicine Group LLC 1; 2018. p. 246–55.

8. Liberati A, Altman DG, Tetzlaff J, Mulrow C, Gøtzsche PC, Ioannidis JPA, et al. The PRISMA Statement for Reporting Systematic Reviews and Meta-Analyses of Studies That Evaluate Health Care Interventions: Explanation and Elaboration. PLoS Med. 2009 Jul 21;6(7):e1000100.

9. Fliefel R, Kühnisch J, Ehrenfeld M, Otto S. Gene Therapy for Bone Defects in Oral and Maxillofacial Surgery: A Systematic Review and Meta-Analysis of Animal Studies. Vol. 26, Stem Cells and Development. Mary Ann Liebert Inc.; 2017. p. 215–30.

10. Sena E, van der Worp HB, Howells D, Macleod M. How can we improve the preclinical development of drugs for stroke? Trends Neurosci. 2007 Sep;30(9):433–9.

